# Association between discrimination stress and suicidality in preadolescent children

**DOI:** 10.1101/2021.05.30.21258084

**Authors:** Stirling T. Argabright, Elina Visoki, Tyler M. Moore, Dallas T. Ryan, Grace E. DiDomenico, Wanjiku F.M. Njoroge, Jerome H. Taylor, Sinan Guloksuz, Ruben C. Gur, Raquel E. Gur, Tami D. Benton, Ran Barzilay

**Author notes:** **Corresponding Author**: Ran Barzilay, MD, PhD, 10th floor, Gates Pavilion, Hospital of the University of Pennsylvania, 34th and Spruce Street, Philadelphia, PA 19104.

## Abstract

**Objective:** US youth suicide rates are increasing in recent years, especially in Black Americans, the reasons for which are unclear. Environmental adversity is key in youth suicidality, hence there is a need to study stressors that disproportionately impact Black youths. We aimed to disentangle the unique contribution of racial/ethnic discrimination from other adversities associated with childhood suicidal ideation and attempts (suicidality) across races.

**Method:** We analyzed data from the Adolescent Brain Cognitive Development (ABCD) Study® that included a large, diverse sample of US children (N=11,235, mean age 10.9 years, 20.2% Black) phenotyped for environmental adversities including discrimination. Multivariate regression models tested the association of self-reported racial/ethnic discrimination with suicidality, covarying for multiple confounders including other discrimination types (towards non-US born individuals; sexual orientation-based; weight-based). Matched analyses contrasted effects of racial/ethnic discrimination and racial identity on suicidality.

**Results:** Black youths reported more discrimination and higher suicidality rates than non-Black youths. High racial/ethnic discrimination was positively associated with suicidality, adjusting for other discrimination types (odds ratio [OR]=2.6, 95%CI=2.1-3.2). Findings remained significant after adjusting for multiple suicidality risk factors. Once experienced, racial/ethnic discrimination was similarly associated with suicidality in White, Black, and Hispanic youths. Matched analyses revealed that racial/ethnic discrimination was associated with suicidality (relative risk [RR]=2.7, 95%CI=2-3.5), while Black race was not (RR=0.9, 95%CI=0.7-1.2).

**Conclusion:** Racial/ethnic discrimination is disproportionately experienced by Black children, and is associated with preadolescent suicidality, over and above other adversities. Findings highlight the need to address discrimination as part of suicide prevention strategies. Cross-sectional design hampers causal inferences.

## Introduction

Suicide rates have seen an upward trend among youths in recent years, roughly doubling between 2007 and 2017.^1–3^ In 2019, suicide was the second leading cause of death among American adolescents aged 13-18, and the fifth leading cause of death in preadolescents aged 6-12.^4^ Black American children are disproportionately affected in the younger age group, dying by suicide at a rate roughly double that of their White counterparts, with many potential contributors including disproportionate exposure to violence, earlier onset of puberty, and diminished access to mental healthcare.^5–7^ To understand this racial imbalance in suicide deaths, there is a need to study specific stressors that disproportionately impact Black American preadolescents in the United States, such as the experience of racial/ethnic discrimination.

Racial/ethnic discrimination – poor and unfair treatment due to one’s race (groups of people with shared physical characteristics often associated with customs/origin) or ethnicity (groups of people with shared customs/origin)^8^ – is a social product of racism that can be experienced in multiple facets of life and may underlie the significant racial health disparities across the lifespan for many minority groups.^7,9,10^ It is known that Black Americans experience significant amounts of racial/ethnic discrimination across the lifespan,^11,12^ which has consistently been shown to associate with poor health outcomes.^13–17^ Preliminary research has extended this work to mental health and suicidal ideation,^18–20^ with similar effects observed across minority groups.^21^ However, the specific role of racial/ethnic discrimination in suicidal ideation or attempts (i.e. suicidality) in children, especially preadolescents, has yet to be established strongly and independently from other environmental adversities.

Determining the unique contribution of racial/ethnic discrimination to suicidality in youth is difficult for many reasons. First, racial/ethnic discrimination is highly correlated with other environmental adversities that negatively impact mental health, such as poverty, trauma, family conflict, and other hardships.^9,22–26^ This web of intertwined environmental exposures, both specific (i.e. childhood trauma) and general (i.e. neighborhood environment), composes the *exposome*, which has major implications for both mental and physical health.^27–29^ Second, racial/ethnic discrimination may be associated with other forms of discrimination related to xenophobia;^30^ sexual orientation;^31^ and weight,^32^ complicating the ability to tease apart the effect of racial/ethnic discrimination on suicidality from that of other discrimination types. Lastly, racial/ethnic discrimination is associated with other (non-suicide-specific) psychopathology, including depression, anxiety, post-traumatic stress, and substance use disorders,^19,33^ which are risk factors for childhood suicidality.^34^ Hence, identifying the specific effects of racial/ethnic discrimination on childhood suicidality requires multiple measures that include deep phenotyping of children’s environment and psychopathology.

Here we sought to disentangle the specific association of racial/ethnic discrimination from other environmental adversities with childhood suicidality in a large, diverse sample of US children from the Adolescent Brain and Cognitive Development (ABCD) Study®.^35^ We aimed (1) to assess the prevalence of self-reported racial/ethnic discrimination in children in the ABCD Study®; (2) to assess the association of racial/ethnic discrimination with suicidality over and above other forms of discrimination (towards non-US born individuals; sexual orientation-based; weight-based) and other adverse exposome factors; and (3) to assess if and how the association between racial/ethnic discrimination and suicidality varies among children of different races or ethnicities. We hypothesized that racial/ethnic minority groups experience a disproportionate amount of racial/ethnic discrimination, and that this experience contributes to childhood suicidality.

## Method

### Participants

The ABCD Study® sample includes 11,878 children aged 9–10 years at baseline, recruited through school systems.^36^ Participants were enrolled at 21 sites, with the catchment area encompassing over 20% of the entire US population in this age group. We included data from ABCD Study® data release 3.0 (https://abcdstudy.org/). In the current analysis we used data collected at the 1-year follow-up assessment (N=11,235, mean age 10.9 years, 52.3% male, 20.2% Black), which included a tool to assess discrimination. In some cases, variables were only available from baseline assessment, either because measures were deemed non-longitudinal (as with race), or because the ABCD data collection schedule did not include these items at the 1-year follow-up (as of ABCD Study® data release 3.0, see **supplemental Table S1** for details on all ABCD measures used in the current study). All participants gave assent. Parents/caregivers signed informed consent. The ABCD protocol was approved by the University of California, San Diego Institutional Review Board (IRB), and was exempted from a full review by the University of Pennsylvania IRB.

### Exposures

The ABCD Study® assessment included an instrument that evaluated youths’ experiences of discrimination.^37^ The instrument was composed of four yes/no questions regarding subjective feelings of discrimination over the past 12 months in four domains: racial/ethnic/color; towards non-US born individuals (i.e., “child or their family are from another [non-US] country”); sexual orientation-based; and weight-based. Thereafter, participants were administered seven questions rating lifetime experiences of discrimination (each question on a 5-point Likert scale, from almost never (1) to very often (5)). The discrimination instrument and frequency of endorsed items are described in **supplemental Table S2**.

The main exposure variable used in the current study was a binary measure of High/Low discrimination. To generate this measure, we used the mean response of the 7 questions (for participants with at least 4 responses, variable dim_y_ss_mean in the ABCD Study®) and determined High discrimination for each child as reporting at or above the 90th percentile. The 7-item discrimination measure was preferred to the binary measure assessing racial/ethnic/color discrimination mainly because of its capture rate; only 4.2% of participants responded to the binary measure versus 11.7% who scored High on the 7-item measure. Additionally, this 7-item measure has a Cronbach’s Alpha of 0.761, supporting its reliability. It was dichotomized due to a skewed distribution of responses toward low scores – to most accurately capture significant experiences of discrimination, we thought it best to take the top decile of scores (see **supplemental Figure S1** for the distribution among participants for this variable).

To our knowledge, this was the first ABCD Study project to use this discrimination tool, providing no precedent as to how to treat discrimination-related measures in analysis. Although the 7-item discrimination measure does not explicitly use the word *race*, it does employ the concept of ethnicity, defined by ABCD as “groups of people who have the same customs, or origin.” Additionally, this item follows the binary item which intertwines the concept of ethnicity with that of race and color (“have you felt discriminated against: because of your race, ethnicity, or color?”). Finally, racial distribution of responses of both measures are similar, with Black participants reporting roughly 3-fold prevalence of discrimination (21.1% Black vs 8.6% non-Black report High discrimination based on 7-item measure; 10.4% Black vs 3.1% non-Black report discrimination based on binary item). Therefore, we refer to our main self-reported discrimination measure as racial/ethnic discrimination throughout the rest of the paper.

### Outcome measures

The Kiddie-Structured Assessment for Affective Disorders and Schizophrenia for DSM-5 (KSADS-5)^38^ assessed suicidal ideation and attempt (past or current).^34^ Items relating to self-injurious behavior without suicidal intent (NSSI) were not included. As the proportion of suicidal attempts was low, and to avoid multiple testing to mitigate risk of type I error, we grouped together suicidal ideation and attempt, in line with previous analyses.^34^ Thus, suicidal outcomes were collapsed into a single binary measure termed *suicidality*. As previous work in the ABCD Study® and other youth samples showed poor agreement between youth and caregiver on suicidality,^24,39^ we referred only to youth report.

### Covariates

All models included age, sex, race (White, Black, and non-Black minority racial groups [Asian, American Indian, Native Hawaiian, and self-identified “other” race]), Hispanic ethnicity, and parental education. In this manuscript, we refer to race categories as “White” and “Black” in order to be consistent with the wording used by the ABCD Study®, where parents were asked which race they considered their child to be: White, Black/African American. To address confounding effects of other types of discrimination, we included the three non-racial/ethnic discrimination variables (past 12-months discrimination towards non-US born individuals, sexual orientation-based, and weight-based – all binary variables), and the identities against which the above discriminations are experienced (i.e., non-US born, lesbian, gay, bisexual, transgender [LGBT], and obese/underweight [BMI>95^th^ percentile/<5^th^ percentile according to CDC]).^40^ To address confounding effects of other environmental adversities we included the following environmental exposures: experiences of poverty in the household (count); family conflict scale; negative life events (count of experiences); and neighborhood socioeconomic status based on estimates of area deprivation using an index accounting for scaled weighted sum of census-tract variables.^41^ Items addressing experiences of household poverty were included in the ABCD longitudinal demographics survey which, due to high collinearity, we collapsed to form a lumped sum-level variable for our analyses (See **supplemental Figure S2** for a scree plot of the 7 items). To address confounding effects of psychiatric diagnoses, we used KSADS-5-based diagnoses of any externalizing disorder (attention deficit hyperactivity disorder [ADHD], oppositional defiant disorder [ODD], or conduct disorder, all based on parent report), or depression or anxiety (based on youth report).^42,43^ Psychosis spectrum was included using the severity score of the prodromal psychosis scale applied in the ABCD Study®.^44^

### Statistical Analyses

We used the SPSS 26.0 statistical package and R 3.6.1 for our data analysis. Mean (standard deviation [SD]) and frequency (%) were reported for descriptive purposes. Univariate comparisons were made using t-tests for continuous measures and chi-square tests for categorical variables, with false discovery rate (FDR) correction for multiple testing when appropriate. We employed listwise deletion for participants with missing data. Rates of missing values for all variables included in the current study were lower than 3.3%, with the exception of discrimination measures that were higher: racial/ethnic (7.4%), sexual orientation-based (6.7%), and weight-based (4.3%). We used two-tailed tests for all models. Data analysis was conducted between November 2020 and January 2021. Data preprocessing and analyses are detailed at https://github.com/barzilab1/abcd_discrimination.

#### Main analysis

To determine the association of racial/ethnic discrimination with suicidality, we conducted a binary logistic regression model with High/Low racial/ethnic discrimination as the independent variable, and suicidality (binary) as the dependent variable, co-varying for age, sex, race (Black, White, non-Black minority racial groups), Hispanic ethnicity, and parental education. To address the study’s main question of the unique effect of racial/ethnic discrimination on suicidality over and above other types of discrimination, we added to the model the three other discrimination variables (towards non-US born individuals; sexual orientation-based; weight-based) and their associated identities (i.e., non-US born, LGBT, and obese/underweight). This model is referred to below as *main model*. Thereafter, we ran two additional models to address specificity of the findings, given that discrimination was associated with other environmental adversities and with other non-suicide psychopathology (see **supplemental Figure S3** for a correlation matrix between discrimination, exposome, and psychopathology). The first model addressed the specificity of racial/ethnic discrimination’s effect on suicidality controlling for multiple other (non-discrimination) environmental adversities (poverty, negative life events, family conflict, and neighborhood deprivation). The second model assessed the direct association between racial/ethnic discrimination and suicidality over and above other (non-suicide) psychopathology through inclusion of variables representing different domains of psychopathology (externalizing and internalizing disorders and psychosis spectrum).

To explore potential differences among races or ethnicities in the magnitude of association between racial/ethnic discrimination and suicidality, we ran the main model stratifying the population to non-Hispanic White, non-Hispanic Black, and Hispanic youths.

#### Sensitivity analyses

We conducted several sensitivity analyses on the main model. To evaluate the potential effect of the method we chose to determine the main exposure (i.e., High racial/ethnic discrimination), we ran the main model substituting the High/Low discrimination variable with the continuous measure averaging all 7 items included in the racial/ethnic discrimination scale. To evaluate the effect of the past 12 months’ experience of racial/ethnic discrimination, we ran the main model substituting the High/Low discrimination variable with the binary yes/no question on the past 12 months’ experience of racial/ethnic/color discrimination. To further address the specificity of racial/ethnic discrimination to suicidality, we ran a combined model that covaried for other discrimination, exposome, and psychopathology measures together. To account for potential effects of family relatedness, we ran two models: one that analyzed data including only one participant (the oldest) from each family and another that excluded families with multiple children entirely.

Lastly, we conducted two sets of matched comparisons that allowed us to contrast the effect of racial/ethnic discrimination with the effect of race on suicidality. In the first matched comparison, we matched High racial/ethnic discrimination to Low racial/ethnic discrimination participants on all measures of the main model, including race. In the second comparison, we matched Black participants to White participants on all measures of the main model, including racial/ethnic discrimination. In both comparisons we used relative risk of suicidality as the outcome variable.

## Results

### Experiences of discrimination and suicidality across races and ethnicity

We first compared the rates of self-reported discrimination in the past 12 months between Black and non-Black participants (**Figure 1A**). Black participants reported over 3-fold more racial/ethnic discrimination compared to non-Black participants (10.4% vs. 3.1%, respectively). In addition, Black participants reported more of the other discrimination types: towards non-US born individuals (2.6% vs. 1.4%), sexual orientation-based (5.8% vs. 3.4%), and weight-based (9.8% vs. 5.1%). Comparison between Hispanic and non-Hispanic participants (**Figure 1B**) revealed similar rates of racial/ethnic discrimination (5.5% vs. 4.4%, respectively) and sexual orientation-based discrimination (3.7% vs. 4%), with higher rates of discrimination towards non-US born individuals (3.9% vs. 1.1%) and based on weight (7.6% vs. 5.7%) experienced by Hispanic participants. Statistics for all above comparisons are presented in **supplemental Table S3**.

**Figure 1.**
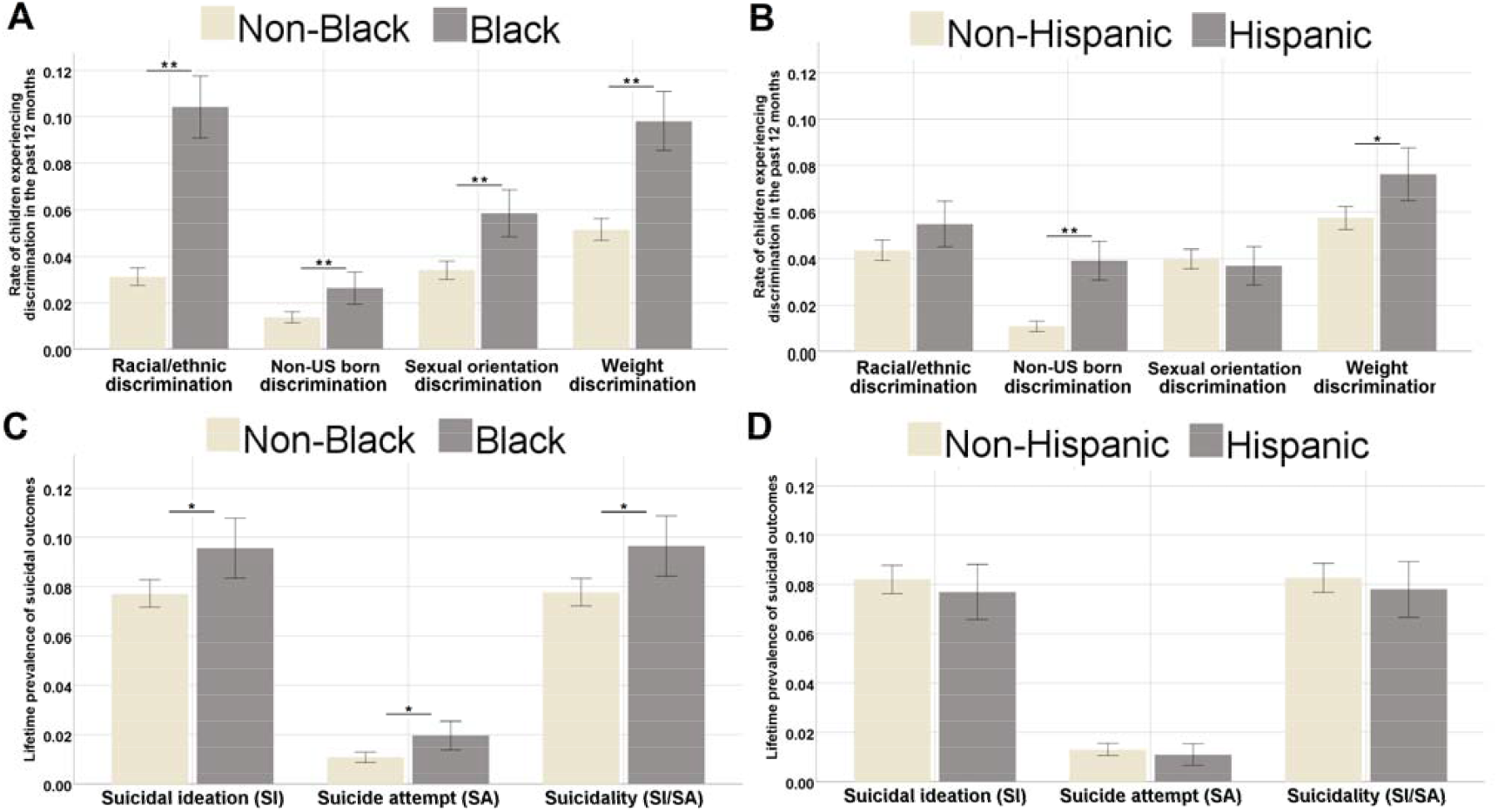
Experiences of discrimination and suicidality across race/ethnicity. Rates of endorsed discrimination and suicidality (proportion 0-1) are displayed across race/ethnicity. **A**. Black participants were compared to non-Black participants in experiences of 4 types of discrimination: racial/ethnic, towards non-US born individuals, sexual orientation-based, weight-based. Black participants experienced significantly higher rates of all 4 types (for all comparisons, FDR corrected *p*-values<0.001). **B**. Hispanic participants were compared to non-Hispanic participants in experiences of 4 types of discrimination: racial/ethnic, towards non-US born individuals, sexual orientation-based, weight-based. Hispanic participants experienced significantly higher rates of discrimination towards non-US born individuals (FDR corrected *p*-value<0.001) and weight-based discrimination (FDR corrected *p*-value=0.004). **C**. Black participants were compared to non-Black participants in experiences of suicidal ideation, suicide attempt, and overall suicidality. Black participants experienced significantly higher rates of all suicidality markers (all FDR corrected *p*-values<0.005). **D**. Hispanic participants were compared to non-Hispanic participants in experiences of suicidal ideation, suicide attempt, and overall suicidality. No significant differences were found (all FDR corrected *p*-values=0.548). \**p*<0.005, \*\**p*<0.001.

Suicidality rates also showed differences across race, as Black participants reported more suicidal ideation and suicide attempts, with 9.7% of Black participants endorsing suicidality compared to 7.8% of non-Black participants (*X*^2^ [1, n=11,077] = 8.399, *p*=0.004; **Figure 1C** and **supplemental Table S3**). No difference in suicidality was found between Hispanic (7.8%) and non-Hispanic (8.3%) participants in suicidality (*X*^2^ [1, n=10,940] = 0.522, *p*=0.548; **Figure 1D** and **supplemental Table S3**).

### Comparison of youths reporting High versus Low racial/ethnic discrimination

We next compared participants who reported High levels (>=90th percentile) of racial/ethnic discrimination to those who reported Low levels (<90th percentile) of racial/ethnic discrimination. Youths endorsing High racial/ethnic discrimination were significantly different from those endorsing Low racial/ethnic discrimination on multiple demographic measures including higher prevalence of males (62% vs. 51%), Black race (38.7% vs. 17.4%), and lower parental education. Furthermore, when compared to Low racial/ethnic discrimination youths, High racial/ethnic discrimination youths reported more discrimination of other types and were generally more likely to experience multiple other adversities including family poverty, negative life events, family conflict, and neighborhood poverty. Lastly, High racial/ethnic discrimination youths had more psychopathology compared to Low racial/ethnic discrimination youths in all diagnostic domains including externalizing and internalizing diagnoses as well as higher psychosis spectrum symptoms and suicidality (19.4% vs. 6.6%, respectively). Full statistics of univariate comparisons are presented in **Table 1**.

**Table 1.**
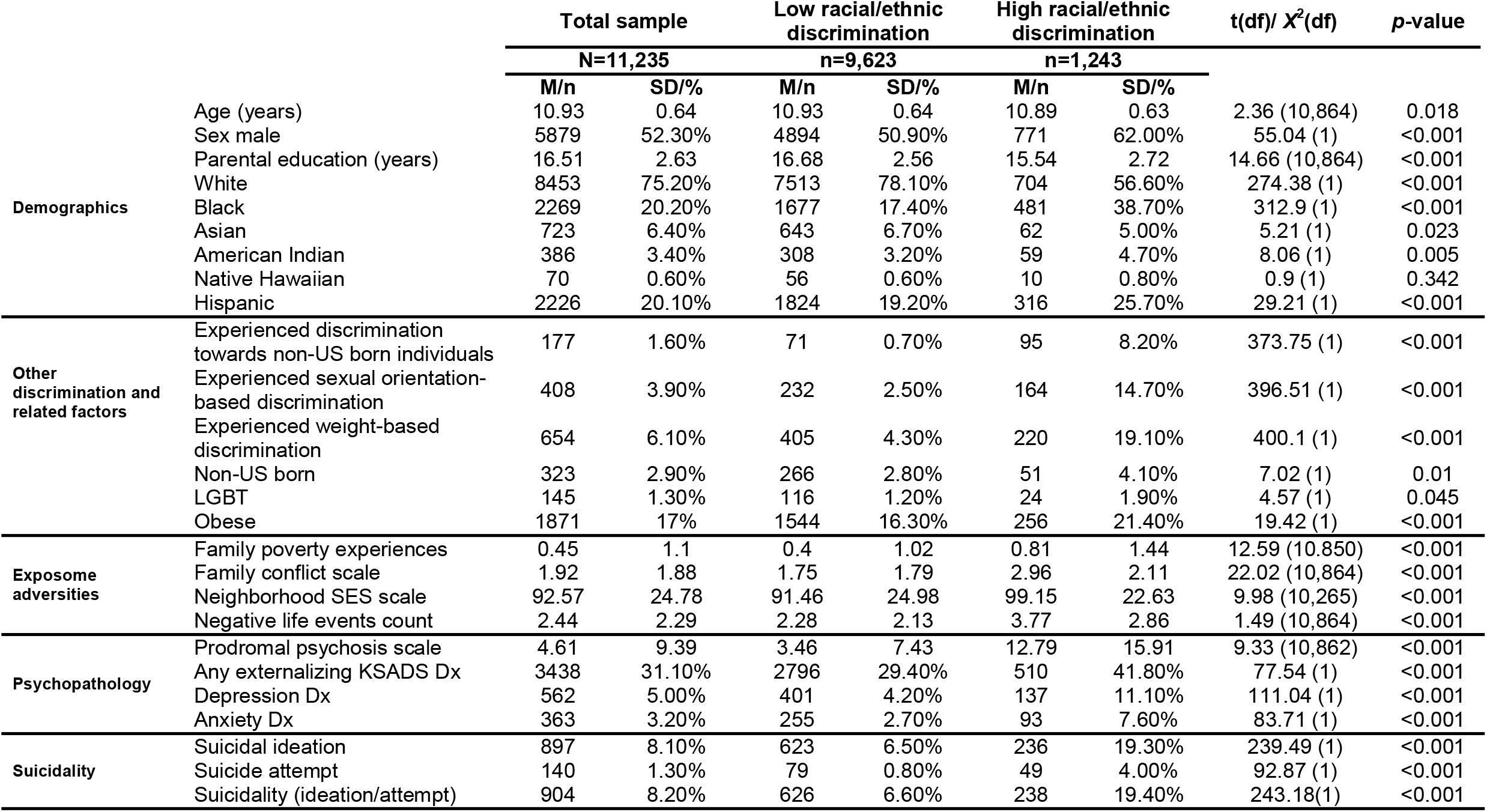
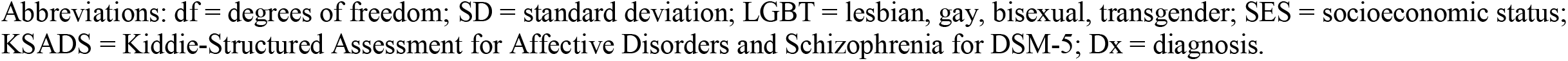
Comparison of youths reporting High versus Low racial/ethnic discrimination When compared across multiple demographic, discrimination-related, environmental exposures, psychopathology, and suicidal behavior, youths experiencing High racial/ethnic discrimination were significantly different from those experiencing Low racial/ethnic discrimination.

### Multivariable modeling

We next sought to delineate the association between High racial/ethnic discrimination and suicidality. High racial/ethnic discrimination was strongly associated with suicidality (odds ratio [OR]=3.5, 95% confidence interval [CI] 2.95-4.16, model co-varied for age, sex, race, ethnicity, and parental education). Our main model revealed that High racial/ethnic discrimination was strongly associated with suicidality (OR=2.6, 95%CI 2.1-3.21) even after co-varying for the other types of discrimination and for the factors based on which this discrimination is experienced (non-US born, identifying as LGBT, and being obese/underweight). The association between High racial/ethnic discrimination and suicidality was robust (OR 1.8, 95%CI 1.43-2.26) to adding multiple environmental adversities to the main model including poverty, negative life events, family conflict, and neighborhood deprivation.

Furthermore, the association was also robust to addition of multiple indicators of psychopathology such as externalizing and internalizing disorders and psychotic symptoms to the main model (OR=1.55, 95%CI 1.22-1.96). Full models’ statistics are detailed in **Table 2**.

**Table 2.**
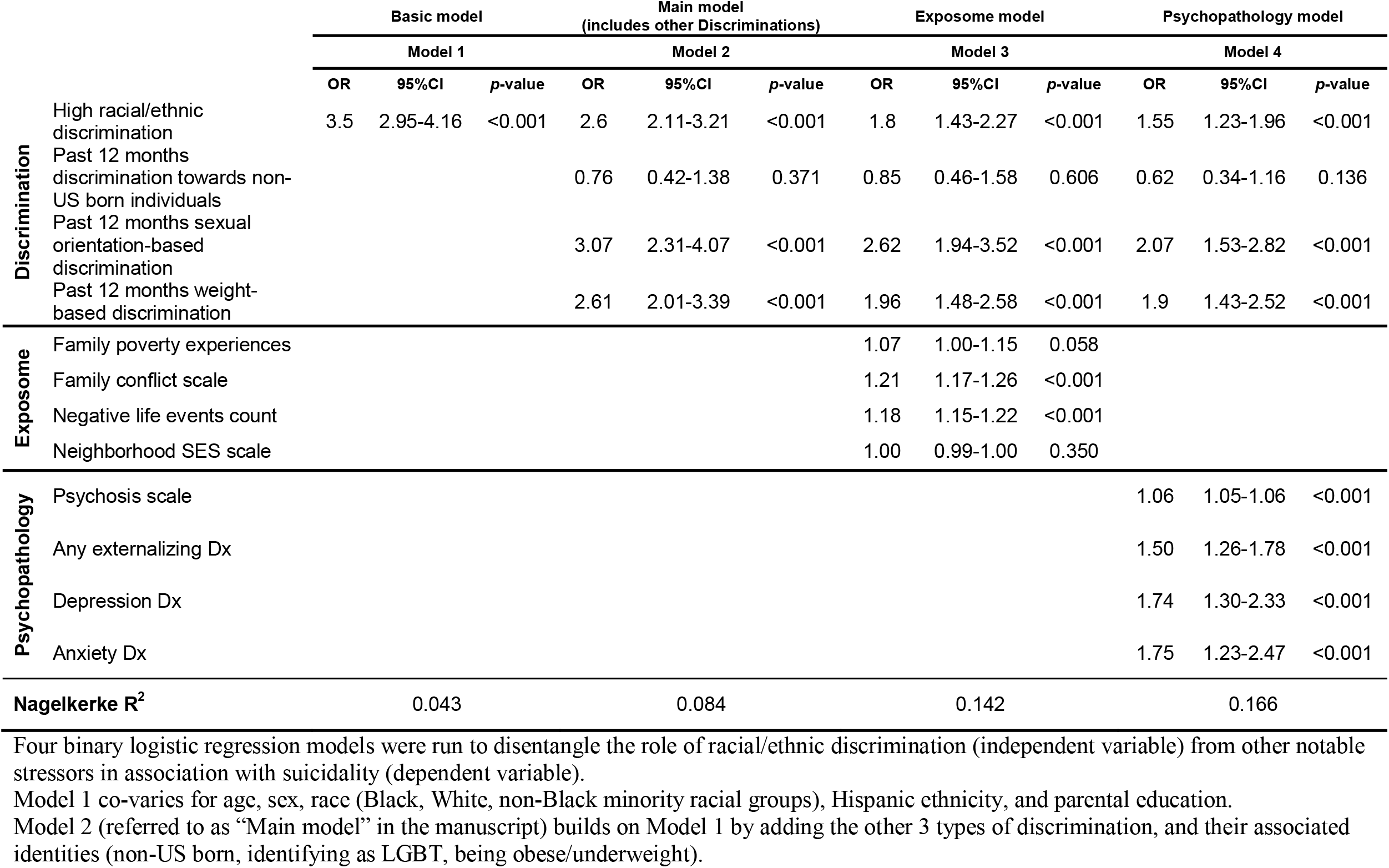
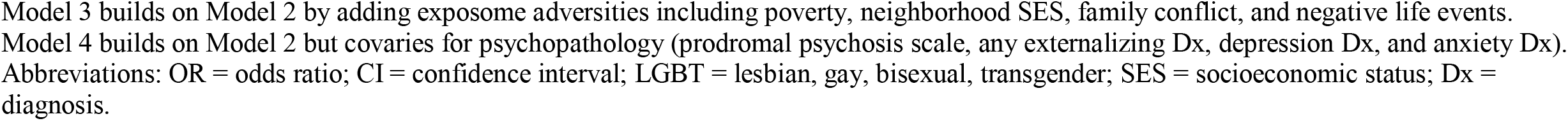
Multivariable modeling of racial/ethnic discrimination’s association with suicidality

### Race/ethnicity stratified analysis

We next explored whether the specific association of High racial/ethnic discrimination (over and above other discrimination types) with suicidality differed among races and ethnicities (**Table 3**). We ran the main model controlling for other discrimination types and found that High racial/ethnic discrimination had similarly deleterious associations with suicidality in non-Hispanic White (OR= 2.96, 95%CI 2.21-3.97), non-Hispanic Black (OR=2.46, 95%CI 1.67-3.61), and Hispanic youths (OR=2.41, 95%CI 1.52-3.83). Effects of High discrimination on suicidality were similar in direction for Asian (OR=1.71, *p*=0.334) and American Indian (OR=1.87, *p*=0.226) youths, though nonsignificant. The Native Hawaiian sample in ABCD (n=70) was underpowered to handle the number of variables used to test the study’s questions in a stratified analysis (**Supplementary Table S4**).

**Table 3.** Race/ethnicity stratified analysis of the main model controlling for other types of discrimination (non-racial/ethnic)

|  | **Non-Hispanic White (n=6,829)** |  |  | **Non-Hispanic Black (n=2,056)** |  |  | **Hispanic youths (n=2,226)** |  |  |
| --- | --- | --- | --- | --- | --- | --- | --- | --- | --- |
|  | **OR** | **95%CI** | **p-value** | **OR** | **95%CI** | **p-value** | **OR** | **95%CI** | **p-value** |
| **High racial/ethnic discrimination** | 2.97 | 2.22-3.98 | <0.001 | 2.46 | 1.67-3.61 | <0.001 | 2.4 | 1.51-3.82 | <0.001 |
| **Past 12 months discrimination towards non-US born individuals** | 0.18 | 0.02-1.42 | 0.103 | 1.62 | 0.58-4.54 | 0.357 | 0.85 | 0.36-2.03 | 0.713 |
| **Past 12 months sexual orientation-based discrimination** | 3.16 | 2.19-4.58 | <0.001 | 2.77 | 1.58-4.86 | <0.001 | 2.65 | 1.30-5.42 | 0.007 |
| **Past 12 months weight-based discrimination** | 2.78 | 1.97-3.94 | <0.001 | 1.84 | 1.06-3.20 | 0.03 | 2.96 | 1.68-5.22 | <0.001 |

Binary logistic regression model with racial/ethnic discrimination as independent variable and suicidality as dependent variable, co-varying for age, sex, race (Black, White, non-Black minority racial groups), Hispanic ethnicity, parental education and the other discrimination types and their associated identities (non-US born, identifying as LGBT, being obese/underweight). Model was run separately on non-Hispanic White, non-Hispanic Black, and Hispanic youths.
Abbreviations: OR = odds ratio; CI = confidence interval; LGBT = lesbian, gay, bisexual, transgender.

### Sensitivity analyses

To address the possibility that our choice of variables influenced the results, we ran the main model using different definitions of racial/ethnic discrimination. Directionality and significance of findings persisted when we used a continuous (rather than our dichotomous High/Low) score of the racial/ethnic discrimination scale (**supplemental Table S5**). Findings remained similar, though nonsignificant (p=0.053), when covarying for other discrimination types, when we used the single binary variable of past 12 months’ racial/ethnic/color discrimination instead of using the 7-item discrimination measure (**supplemental Table S6**). Sensitivity analyses addressing within-family effects yielded similar directionality and significance as well (**supplemental Tables S7** and **S8**). When covarying for other discrimination, exposome, and psychopathology measures together in a combined model, results were similar in direction but nonsignificant (*p*=0.092).

### Matched comparisons

High discrimination was associated with increased risk of suicidality (relative risk [RR]= 2.65, 95%CI 2.02-3.47, **Table 4**) when comparing High racial/ethnic discrimination youths to Low racial/ethnic discrimination youths matched on multiple measures including age, sex, race, and other discrimination types (n=954 in each group, for characteristics of matched sample see **supplemental Table S9**). In contrast, Black race was not associated with suicidality (RR=0.94, 95%CI 0.74-1.19) when comparing Black to White youths matched on levels of racial/ethnic discrimination, age, sex, and other discrimination types (n=1,399 in each group, **supplemental Table S9**).

**Table 4.** Matched analyses

|  | Matched analysis 1 |  |  |  |  |  |  | Matched analysis 2 |  |  |  |  |  |  |
| --- | --- | --- | --- | --- | --- | --- | --- | --- | --- | --- | --- | --- | --- | --- |
|  | Low racial/ethnic discrimination<br>(n=954) |  | High racial/ethnic discrimination<br>(n=954) |  | RR | 95%CI | p-value | White<br>(n=1,399) |  | Black<br>(n=1,399) |  | RR | 95%CI | p-value |
|  | n | % | n | % |  |  |  | n | % | n | % |  |  |  |
| **Suicidality** | 65 | 6.90% | 172 | 18.30% | 2.65 | 2.02-3.47 | <0.001 | 130 | 9.40% | 122 | 8.80% | 0.94 | 0.74-1.19 | 0.528 |

In matched analysis 1, participants reporting High racial/ethnic discrimination were matched with participants reporting Low racial/ethnic discrimination on multiple measures including age, sex, race, and all other discrimination types to participants.
In matched analysis 2, Black American participants were matched to White American participants on levels of racial/ethnic discrimination, age, sex, and other discrimination types.
Abbreviations: RR = relative risk; CI = confidence interval.

## Discussion

In this study we investigated the association of racial/ethnic discrimination with preadolescent suicidality. In line with the recent literature, we observed that Black American youths in the ABCD Study® report higher levels of racial/ethnic discrimination and display more suicidality than other racial groups.^9,11^ Notably, we found that Black children experience disproportionate amounts of *all* studied forms of discrimination (towards non-US born individuals; sexual orientation-based; weight-based) compared to non-Black children – a finding that, to the best of our knowledge, has not yet been documented in a large sample of American children. Multivariate analyses revealed that racial/ethnic discrimination is a unique contributor to suicidality in American youth, independent of other known environmental risk factors for suicidality including negative life events,^23^ family conflict,^34^ other discrimination types (i.e. sexual orientation-based),^31^ and psychopathology.^25,45^ Furthermore, though racial/ethnic discrimination is disproportionately experienced by Black children, its association with suicidality is similar in non-Black children. This association became nonsignificant when Asian, American Indian, and Native Hawaiian participants were examined independently, though this is likely due to insufficient power due to their smaller sample size among ABCD Study’s participants. These findings suggest that racial/ethnic discrimination is a major stressor uniquely associated with preadolescent suicidality.

The assessment of discrimination in the ABCD Study® included items related to four different domains: racial/ethnic; towards non-US born individuals; sexual orientation-based; and weight-based. Our main analysis examined the association of racial/ethnic discrimination with suicidality, controlling for key demographic factors as well as other forms of discrimination and their associated identities (i.e. non-US born, LGBT, obese/underweight). We found that racial/ethnic discrimination imposes a unique psychological stress that is significantly associated with childhood suicidality with a magnitude of effect similar to well established risk factors like sexual orientation-based discrimination^31^ and weight-based discrimination.^32^ Notably, the data analyzed in the current study was collected in 2018, two years before the COVID-19 pandemic, post-election furor, and the prominent racial justice protests of 2020-2021; therefore, our findings may have been even more pronounced had the data been collected today, in 2021.

A fundamental challenge of studying environmental effects on health outcomes is that they are often co-linear, and it is difficult to disentangle the unique effect of any specific stressor. The exposome framework embraces the inherent complexity of the environmental network rather than focusing on a single adversity (e.g., trauma), examining environmental exposures within dynamic interactive domains.^28,29^ The large sample size and deep phenotyping of the ABCD Study® cohort provides an opportunity to dissect the specific components of the exposome and their interactions with race. Such has been extensively studied by Assari et. al., who has highlighted racial discrepancies in the effects of environmental protective factors.^46,47^ Similarly, we sought to test the association of racial/ethnic discrimination on suicidality within the scope of the exposome. Discrimination was significantly correlated with adverse exposome and psychopathology measures, both of which are documented risk factors that may inflate its effect size on suicidality. However, racial/ethnic discrimination remained strongly associated with suicidality even when accounting for environmental adversity – poverty, low socioeconomic status, negative life events, family conflict^9,22,24,26^ – and non-suicide psychopathology, represented by diagnosis of both internalizing and externalizing domains.^25,45^ It is important to note that items related to NSSI were not included. Although NSSI and suicidality are highly related, there is evidence that the two have distinct etiologies and the relationship between the two is complex and beyond the scope of this analysis.^48^ Regardless, results highlight the burden that racial/ethnic discrimination has on mental health in childhood, demonstrating its effect over and above other environmental exposures.

A major finding that we report here is that once racial/ethnic discrimination is experienced, it seems to retain its deleterious association with suicidality across races and ethnicities (i.e., in both White and Black children; in both Hispanic and non-Hispanic children). Our matched analyses allowed us to inspect the role of discrimination versus that of race in association with suicidality. When High discrimination participants were matched to Low discrimination participants at every other variable, we found that High racial/ethnic discrimination was robustly associated with elevated levels of suicidality. However, when Black participants were matched to White participants at every other variable, there was no significant association between Black race and suicidality. Our findings resonate with a recent study by Matheson et. al., which found consistency in the effect of discrimination on mental health in various groups that have been historically marginalized, including Indigenous peoples, Black individuals, Jewish individuals, and women.^21^

Our findings have some immediate implications for clinicians and suicide researchers. We demonstrate that experiences of racial/ethnic discrimination are significant stressors whose magnitude of association with increased suicidality is similar to that of well-established risk factors, such as sexual orientation-based discrimination^31^ or history of depression.^45^ Therefore, clinicians should be mindful of this unique stressor and consider this as a potential contributor to suicide risk. Notably, if a clinician decides it is worthwhile to bring up racial/ethnic discrimination in a clinical evaluation, it should not be limited to Black children or children of other minority groups; the results attest that non-Black children who feel racially/ethnically discriminated against also have a higher chance of endorsing suicidality compared to their counterparts who do not feel discriminated against. It is important, however, that any discussion regarding race/ethnicity should be done with care to avoid further mental anguish, as racial issues can be mentally burdensome, potentially traumatic topics for affected children if not handled with sensitivity.^12,49^

The current findings should be interpreted in the context of several limitations. First and foremost, it is important to address the limitations with the measures used to assess racial/ethnic discrimination. In the 7-item discrimination measure used in main analyses, only the first 4 items refer specifically to ethnicity (defined by ABCD as “groups of people who have the same customs, or origin”), while the latter 3 items focus on feelings of marginalization/ostracization. This is in contrast to the binary variable assessing past 12-months’ experiences of racial/ethnic/color discrimination. However, although these concepts are nuanced and not perfectly equivalent, they are heavily entangled, and it is reasonable to assume they were perceived by the child participants as strongly associated. This is reflected in the similar distributions of Black versus non-Black participants endorsing High racial/ethnic discrimination based on the 7-item measure (21.1% vs 8.6%) and racial/ethnic/color discrimination in the past 12 months (10.4% vs 3.1%). Additionally, because racial/ethnic discrimination was measured using a 7-item questionnaire while other forms of discrimination were measured using a binary question regarding the past 12 months, the effect of racial/ethnic discrimination may be inflated over other discrimination types. Similarly, covariate identities based on assessed discrimination types were not exactly aligned. For example, the identity associated with discrimination towards non-US born individuals was based only on whether or not the child was US-born, excluding first generation Americans who may experience this same form of discrimination, and the identity associated with sexual discrimination included transgender individuals, an identity that was not mentioned in the question on discrimination. Still, we show significant associations of multiple discrimination types with suicidality. We also demonstrate that racial/ethnic discrimination retains its positive significant association with suicidality even when accounting for many confounders in sensitivity analyses, suggesting that despite the limitations above, our findings add a timely and important perspective on the factors contributing to the growing problem of preadolescent suicidality.

Another concern is the personal characteristics of the High racial/ethnic discrimination sample. One may argue that the observed association between discrimination and suicidality may be inflated by poor self-esteem, low socioeconomic status, family neglect, or negative outlook, which may cause a participant to feel more highly discriminated against. We believe that this concern is greatly mitigated by the fact that the discrimination-suicidality association remained significant even following rigorous covariation for demographic and socioeconomic factors, and environmental adversity. The association was also significant in models that accounted for prodromal psychosis symptoms, depression, and anxiety, mitigating the concern that underlying psychopathology explains this discrimination-suicidality association. When other discrimination, exposome, and psychopathology factors were covaried for in the same model, however, findings were nonsignificant. Additionally, heterogeneity within assessed groups, especially “White” participants, may skew findings and limit generalizability. For example, self-identified “White” race often includes minority groups (e.g. Middle Eastern descent, Muslim and orthodox Jewish individuals), which may inflate instances of self-reported discrimination.^21,50^ Moreover, participants are recruited from sites across the US, where amount and type of discrimination may vary based on demographic composition of the area and associated study population. The study’s cross-sectional design also limits causal inferences between self-reported discrimination and suicidality. Nonetheless, the rigorous inclusion of confounders often associated with racial/ethnic discrimination and the matched analyses underscoring its unique effect support the directionality of the association from discrimination to suicidality. Future longitudinal studies are needed to clarify causal pathways in this cohort and others.

In conclusion, we report that the subjective experience of racial/ethnic discrimination is robustly and independently associated with suicidality in children, regardless of race or ethnicity. Findings suggest that race itself is not associated with suicidality, but rather the experience of discrimination associated with belonging to a minority racial/ethnic group in the United States is the factor that contributes to suicidality. Black American children, however, are disproportionately exposed to racial/ethnic discrimination and therefore bear a disproportionate psychological burden. These findings may imply that care providers should be aware of discrimination’s detrimental effect on childhood mental health. Finally, while we focus on a US sample, such discrimination is likely present in other countries with multi-racial/ethnic populations, which merits further investigation globally.

## Data Availability

Data used in the preparation of this article were obtained from the Adolescent Brain Cognitive DevelopmentSM (ABCD) Study (https://abcdstudy.org), held in the NIMH Data Archive (NDA).

## Acknowledgments

This study was supported by the National Institute of Mental Health grants K23MH120437 (RB), R21MH123916 (RB), RO1MH117014 (TMM and RCG) and the Lifespan Brain Institute of Children’s Hospital of Philadelphia and Penn Medicine, University of Pennsylvania.

Data used in the preparation of this article were obtained from the Adolescent Brain Cognitive Development^SM^ (ABCD) Study (https://abcdstudy.org), held in the NIMH Data Archive (NDA). This is a multisite, longitudinal study designed to recruit more than 10,000 children age 9-10 and follow them over 10 years into early adulthood. The ABCD Study® is supported by the National Institutes of Health and additional federal partners under award numbers U01DA041048, U01DA050989, U01DA051016, U01DA041022, U01DA051018, U01DA051037, U01DA050987, U01DA041174, U01DA041106, U01DA041117, U01DA041028, U01DA041134, U01DA050988, U01DA051039, U01DA041156, U01DA041025, U01DA041120, U01DA051038, U01DA041148, U01DA041093, U01DA041089, U24DA041123, U24DA041147. A full list of supporters is available at https://abcdstudy.org/federal-partners.html. A listing of participating sites and a complete listing of the study investigators can be found at https://abcdstudy.org/consortium_members/. ABCD consortium investigators designed and implemented the study and/or provided data but did not necessarily participate in analysis or writing of this report. This manuscript reflects the views of the authors and may not reflect the opinions or views of the NIH or ABCD consortium investigators.

